# A single night in hypoxia either with or without ketone ester ingestion reduces sleep quality without impacting next day exercise performance

**DOI:** 10.1101/2024.08.20.24311732

**Authors:** Myrthe Stalmans, Domen Tominec, Robberechts Ruben, Wout Lauriks, Monique Ramaekers, Tadej Debevec, Chiel Poffé

**Affiliations:** Exercise Physiology Research Group, Department of Movement Sciences, KU Leuven, Leuven, Belgium; Faculty of Sport, University of Ljubljana, Ljubljana, Slovenia; Department of Automatics, Biocybernetics and Robotics, Jožef Stefan Institute, Ljubljana, Slovenia; REVAL – Rehabilitation Research Center, Faculty of Rehabilitation Sciences, Hasselt University, Diepenbeek, Belgium

**Keywords:** ketone, sleep, recovery, oxygen saturation, exercise performance, hypoxia

## Abstract

**Background:** Sleeping at (simulated) altitude is highly common in athletes as an integral part of altitude training camps or sport competitions. However, it is also often feared due to proclaimed negative effects on sleep quality, thereby potentially hampering exercise recovery and next-day exercise performance. We recently showed that ketone ester (KE) ingestion beneficially impacted sleep following strenuous, late evening exercise in normoxia, and alleviated hypoxemia. Therefore, we hypothesized that KE ingestion may be an effective strategy to attenuate hypox(em)ia-induced sleep dysregulations.

**Methods:** Eleven healthy, male participants completed three experimental sessions including normoxic training and subsequent sleep in normoxia or at a simulated altitude of 3,000m while receiving either KE or placebo post-exercise and pre-sleep. Sleep was evaluated using polysomnography, while next-day exercise performance was assessed through a 30-min all-out time trial (TT_30’_). Physiological measurements included oxygen status, heart rate variability, ventilatory parameters, blood acid-base balance and capillary blood gases.

**Results:** Hypoxia caused a ∼3% drop in sleep efficiency, established through a doubled wakefulness after sleep onset and a ∼22% reduction in slow wave sleep. KE ingestion alleviated the gradual drop in SpO_2_ throughout the first part of the night, but did not alter hypoxia-induced sleep dysregulations. Neither KE, nor nocturnal hypoxia affected TT_30’_ performance, but nocturnal hypoxia hampered heart rate recovery following TT_30’_.

**Conclusion:** We observed that sleeping at 3,000m altitude already impairs sleep efficiency. Although this hypoxia-induced sleep disruption was too subtle to limit exercise performance, we for the first time indicate that sleeping at altitude impairs next-day exercise recovery. KE alleviated nocturnal hypoxemia whenever SpO_2_ values dropped below ∼85%, but this did not translate into improved sleep or next-day exercise performance.

## INTRODUCTION

Sleeping at (simulated) altitude is highly common in athletes as an integral part of altitude training camps. However, athletes are also often enforced to sleep at altitude during major sport competitions such as grand tours in cycling, Olympic Games, multi-day ultrarunning races or mountaineering expeditions. During altitude training camps, nocturnal hypoxic exposure is considered essential for inducing favorable physiological adaptations in the longer term such as increased hemoglobin mass and maximal oxygen consumption rates which may ultimately improve endurance exercise performance (1). Conversely, sleeping at altitude is also often feared by athletes – especially during competitions – due to proclaimed negative effects of hypoxia on sleep quality which may in turn hamper exercise recovery and next-day exercise performance.

In this respect, studies generally observed that a single night at (simulated) altitudes above 2,000 m impairs sleep quality (2–10). This is evidenced by decreases in either rapid eye movement (REM) or slow wave sleep (SWS), or an increase in wakefulness during the night. Methodological heterogeneity and contradictory findings, however, hinder unequivocal conclusions and a clear rationale. In general, REM sleep is unaffected at altitudes up to 3,800m (2, 4, 6, 7, 11–13) but decreases up to 40% at altitudes starting from 4,000m (3, 5, 9, 10). Data on SWS is much more equivocal with some studies reporting no changes at altitudes up to 3,800m (7, 11, 12), while other studies reported decreases from ∼20-45% at altitudes between 2,000 and 4,300m (2, 5, 6, 9, 13). This is generally accompanied by an increased wakefulness after sleep onset (WASO) (3–7, 9). Conversely, sleep efficiency typically remains unaffected by altitudes of up to 4,000m (2–4, 6, 7, 11–13), but decreases by 10-20% at 4,000m (3, 13) and higher (9).

Sleep disturbances may have both direct and indirect implications for athletic performance. This is evidenced by studies showing that a single night of restricted sleep (i.e., sleep time reduction of 2-4h) increases cardiac stress during and after exercise on the next day (14). Furthermore, ∼5 days of disturbed sleep has been shown to suppress myofibrillar and sarcoplasmic protein synthesis rates (15, 16), to impair insulin sensitivity (17), and to reduce mitochondrial respiratory function (16). Although some studies reported no impact of sleep restriction down to 2h per night on next day exercise performance (14, 18), other studies indicated that a single night of ∼2-3h sleep deprivation is sufficient to decrease 6s peak cycling power output and both 3 and 20 km cycling time-trial performance (19, 20). Nevertheless, it is currently unknown if the more subtle differences in sleep quality and quantity that are expected to occur upon a single night in hypoxia also impair next day exercise performance.

The precise underlying mechanisms of hypoxia-induced sleep impairments are currently unknown. Yet, they are most likely caused by hypoxemia (8, 12, 21, 22). Interestingly, we previously showed that increasing blood ketone bodies (KB), via oral ketone ester (KE) ingestion, attenuated the drop in blood (as well as muscle and brain) oxygenation after 3-4h in hypoxia (23, 24). Furthermore, we identified that KE intake post-exercise and pre-sleep beneficially impacted sleep following strenuous, late evening exercise in normoxic conditions. This was evidenced by an inhibition of late evening exercise-induced decreases in REM sleep and WASO, and a 2% improvement in sleep efficiency (25).

These data indicate that pre-sleep KE ingestion may be an effective strategy to attenuate hypox(em)ia-induced sleep dysregulations. Against this background, this study aims to identify the impact of a single night in hypoxia on (i) nocturnal sleep measured using golden standard polysomnography, and (ii) next day exercise performance, and to (iii) explore the potential of KE supplementation to counteract potential performance or sleep dysregulations under these conditions.

## METHODS

### Ethical approval and participants

This research was registered at www.clinicaltrials.gov (NCT06060093) and approved by the Ethics Committee Research UZ/KU Leuven (B3222022001041). Potential participants were medically screened and provided written informed consents. Candidates’ sleep quality was screened using the Pittsburgh Sleep Quality Index (PSQI) in order to guarantee adequate baseline sleep quality as defined by the National Sleep Foundation (i.e., sleep efficiency above 85%). Participants working in late-night shifts, as well as extreme morning and evening chronotypes as determined by the Horne-Östberg questionnaire were excluded. All thirteen healthy, male participants that were included reported good sleep quality and no indications for sleep disorders (PSQI score <5). None of them showed indications for psychological or neurological disorders, such as depression (Beck’s Depression questionnaire) or anxiety (Beck’s Anxiety questionnaire). All participants were active in cycling, endurance running or triathlon. Further exclusion criteria included smoking, exposure to altitudes >1,500m during the 3 months preceding the study or a history of altitude-sensitive pathologies. From the thirteen participants that were initially included, one dropped out due to a Covid-19 infection and another one was excluded from the data analyses due to a baseline sleep quality drastically below the predefined inclusion criteria (i.e., sleep efficiency of 78%). Therefore, data analyses were performed on 11 participants [age: 24 ± 4 yr; height: 181.3 ± 7.5 m; body mass: 73.2 ± 9.2 kg; physical activity: 8 ± 3 h.week^−1^, VCO_2_max: 61.5 ± 8.8 mL.kg^−1^.min^−1^ (mean ± SD)].

### General study design

This randomized, double-blind, placebo-controlled, cross-over study consisted of three experimental sessions. These sessions were designed to mimic a live-high, train-low setup, and all included an identical training session in normoxia whereafter participants either spend the night in (i) normoxia (N_PL_) or (ii) normobaric hypoxia (∼3,000m simulated altitude) while receiving a placebo (H_PL_), or in (iii) hypoxia while receiving KE (H_KE_, Fig. 1). Following the night in either hypoxia or normoxia, participants performed an all-out 30min time-trial (TT_30’_) to evaluate exercise performance. Physiological measurements were performed at rest throughout both days of the protocol. This design allowed us to evaluate both (i) the impact of sleeping in hypoxia *vs.* normoxia, and (ii) the potential of KE to improve sleep in hypoxia. All experimental sessions were conducted in a normobaric hypoxic facility (Van Amerongen CA Technology, Tiel, The Netherlands) at the Bakala Athletic Performance Center (Leuven, Belgium). The experimental sessions were separated by a one-week wash-out period, and experimental protocols and timings were identical for all three sessions.

**Figure 1.**
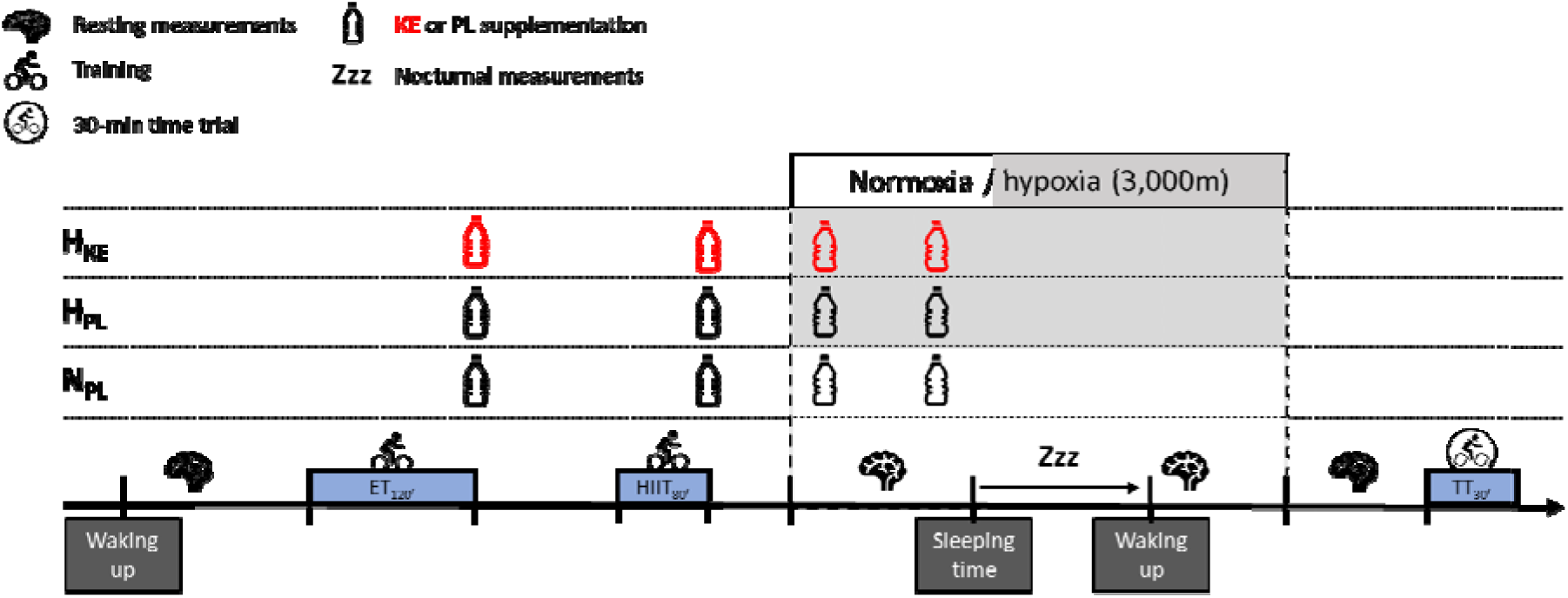
Schematic representation of the experimental protocol that was designed to mimic a live-high train-low (LHTL) strategy. In a double-blind, randomized, crossover design, 11 participants completed three experimental sessions each involving a two-day protocol. On day 1, subjects performed two exercise training sessions [120 min endurance training (ET_120’_) and 80 min high intensity interval training (HIIT_80’_)] in normoxia followed by a night either in normoxia (N), or at a simulated altitude of 3,000m (H). After each training session and before sleep, participants received either placebo (N_PL_ and H_PL_) or ketone ester (H_KE_) supplements. On day 2, exercise performance was evaluated by means of a 30-min all-out time trial (TT_30’_). Sleep quality and architecture were assessed via polysomnography. Physiological measurements were performed in a resting state at baseline, after 2h in hypoxia or equivalent in normoxia, immediately upon waking up and 2h after return to normoxia in order to assess blood oxygen saturation, cerebral and muscular oxygenation status, ventilatory gas exchange, heart rate and heart rate variability. In addition, before every resting measurement, as well as immediately before sleep time, a capillary blood sample was collected for determination of acid-base balance, and blood gasses.

### Preliminary testing and familiarization sessions

Two weeks before the first experimental session, participants performed a familiarization session consisting of a graded exercise test on a cycling ergometer (Avantronic Cyclus II, Leipzig, Germany) to assess both their lactate threshold (LT) and maximal oxygen uptake rate (VCO_2_max). For the LT determination, initial workload was set at 70W and increased by 40W per 8min. Capillary blood samples were obtained every 4min for determination of blood lactate concentration (Lactate Pro2; Arkray, Amstelveen, The Netherlands), with LT being defined as the lowest workload provoking a 1 mM blood [lactate] increase within the same stage (25–27). After 15min of active recovery at 70W, participants started a second incremental cycling test (100W + 25W/30sec) until volitional exhaustion. Throughout this test, oxygen consumption (VCO_2_) and carbon dioxide production (VCCO_2_) rates were measured continuously using indirect calorimetry (Cortex Metalyzer 3B, Cortex, Leipzig, Germany), and VCO_2_max was calculated as the highest VCO_2_ over a 30-sec period.

In the 2 weeks before the first experimental session, participants performed the full experimental protocol twice in normoxia to get accustomed to the experimental procedures, exercise protocols, and sleeping facility. Starting from the first familiarization session, participants were asked to maintain a stable sleep schedule until the final experimental session. This included the requirement to sleep 7-9h per night, with bed-time between 10-12 PM and wake-time between 6-8 AM. Compliance to a stable sleep-wake schedule was assessed throughout the entire study period via a sleep diary and actigraphy (Actigraph wGT3X-BT, ActiGraph LLC, Pensacola, Florida, US). Moreover, sleep quality of the night prior to each experimental session was also assessed using the St. Mary’s Sleep questionnaire.

### Experimental sessions

After waking up at their individually predetermined time, participants arrived in a fasted state between 7.00 and 9.00 AM at the testing facility (exact timings were replicated for each session). 2.5h after consuming a standardized breakfast (see *Dietary standardization* below), they performed a 2-h morning endurance training session (ET_120’_) on a cycling ergometer (Tacx Neo Smart, Wassenaar, The Netherlands). ET_120’_ consisted of eight consecutive 15-min intervals with workload alternating between 60% and 80% of LT. 1.5h after lunch time, participants completed an 80-min high intensity interval training (HIIT_80’_) on a cycling ergometer (Avantronic Cyclus II, Leipzig, Germany). HIIT_80’_ consisted of a 10-min warm up at 70% of LT, followed by 10 repetitions of 3min at 120% of LT interspersed by 4min active recovery at 50% of LT. Thirty minutes after HIIT_80’_ (∼5h before sleep-time), participants entered their individual ‘hotel room’ that was set at either hypoxia (H_PL_ and H_KE_; F_I_O_2_: 14.5% O_2_, P_I_O_2_: ∼110 mmHg, ∼3,000m simulated altitude) or normoxia (N_PL_; F_I_O_2_: 20.9% O_2_, P_I_O_2_: ∼160 mmHg). Subsequently participants resided for 16h under these environmental conditions, with sleep being measured in each condition using polysomnography (PSG). Following 16h of hypoxia/normoxia, participants returned to normoxia and exercise performance was assessed 3.5h later by means of an all-out 30-min time trial (TT_30’_). In addition, physiological measurements were performed at rest at the following timepoints: (i) before ET_120’_ (baseline, 0h), (ii) after 2h in hypoxia or equivalent in normoxia (+10h), (iii) immediately upon waking up (+23h), and (iv) 2h after returning to normoxia (+27h). These measurements included determination of heart rate (HR), heart rate variability (HRV), ventilatory parameters, and blood, brain and skeletal muscle oxygenation.

### Supplementation protocol

In a randomized order, participants received 25g of either a ketone ester [H_KE,_ 96% (R)-3-hydroxybutyl (R)-3-hydroxybutyrate, KetoneAid Inc., Falls Church, Virginia, USA] or a placebo (N_PL_ and H_PL,_ details below) supplement immediately after (i) ET_120’_ and (ii) HIIT_80’_, (iii) after 1.5h in hypoxia (or equivalent in normoxia), and (iv) 30min before sleep. Hence, in H_KE_ participants received a total dose of 100g ketone ester (1.39 ± 0.17 g·kg^−1^ body weight) in order to intermittently elevate blood ketone bodies post-exercise and throughout the first part of the night. In N_PL_ and H_PL_, participants received a total of 100g of a taste and viscosity matched, inert placebo consisting of 12.5% w/v collagen (6d Sports Nutrition, Oudenaarde, Belgium) and 1 mM bitter sucrose octaacetate (Sigma-Aldrich, Bornem, Belgium) dissolved in water. In total, caloric intake via the KE supplements was ∼700 kcal *vs.* ∼0 kcal for the placebo supplements. Randomization was performed by a researcher who was otherwise not involved in the study.

### Dietary standardization

The evening before each experimental session, participants consumed a standardized carbohydrate-rich dinner at home (∼5,600 kJ; 69% carbohydrate, 16% fat, 15% protein). Water consumption was allowed *ad libitum* until arrival at the facility the next morning, and was replicated for each session. After participants arrived at the testing facility in a fasted state, they consumed a standardized breakfast (∼4,200 kJ; 68% carbohydrate, 21% fat, and 11% protein). Subsequently participants received a carbohydrate-rich snack (∼660 kJ; 92% carbohydrate, 3% fat, and 5% protein) 1h before ET_120’_, and received 60g carbohydrates per hour during ET_120’_ via isotonic drinks and carbohydrate-rich snacks (6d Sports Nutrition, Oudenaarde, Belgium). A light lunch (∼4,150 kJ; 74% carbohydrate, 25% fat, and 14% protein) was provided 1.5h before HIIT_80’_, and participants received a high-carbohydrate, high-protein recovery shake (60g carbohydrate, 30g protein, 6d Sports Nutrition, Oudenaarde, Belgium) immediately after HIIT_80’_. After 30min in hypoxia/normoxia, a standardized dinner (∼3,250 kJ; 69% carbohydrate, 5% fat, and 26% protein) was served while participants received a light snack (∼1,700 kJ of which 69 % carbohydrates, 15 % protein, 16 % fat) 2h before sleep time in order to prevent participants from going to bed hungry. Hence, total caloric intake on the first day – excluding the KE supplements – was ∼3,590 kJ. The next morning, participants received an identical breakfast as on the first day, and received a carbohydrate-rich snack (cfr. before ET_120’_) 30min before TT_30’_.

### Nocturnal measurements

#### Polysomnography (PSG)

A digital amplifier (V-amp, Brain Products, Gilching, Germany) was used to record sleep quality and quantity during the experimental sessions, and data were digitized at a sampling rate of 1000 Hz. According to the international 10 – 20 system, electroencephalographic (EEG) recordings were made from Fz, Cz, Pz, Oz, C3, C4, A1, and A2 where the A2 electrode served as the reference electrode and A1 as a backup reference. A ground electrode was positioned in the middle of the forehead. Eye movements (vertical/horizontal) were recorded with electrooculographic (EOG) electrodes above and under the right eye and with electrodes attached to the outer cantus of both eyes. A 0.1 Hz low cut-off filter and a 30 Hz high cut-off filter were applied to both EEG and EOG data. Submental muscle tone and movements were assessed with a electromyogram (EMG) of the chin, recorded with a low cut-off filter of 10 Hz and a high cut-off filter of 200 Hz. Electrical noise was filtered out with a 50 Hz notch filter. A blinded, independent, certified sleep technician (Sleep Well PSG, Canada) visually scored night recordings following the Rechtschaffen & Kales guidelines, in conjunction with the AASM guidelines, in 30 s epochs. A 0.3 – 35 Hz, 0.3 – 30 Hz, and a 10 – 100 Hz bandpass filter was used for EEG, EOG and EMG signals, respectively. The assessed sleep variables were: (i) total sleep time, (ii) WASO, (iii) total NREM and REM sleep, (iv) total N1, N2, and REM and SWS sleep, (v) sleep onset, sleep onset to N2, SWS, and REM sleep, (vi) sleep efficiency (total sleep time/time in bed), and (vii) amount of awakenings.

#### Sleep event detection

EEG data were preprocessed in BrainVision Analyzer (Brain Products GmbH, Gilching, Germany). After applying a 0.1 – 30 Hz bandpass filter, data were transferred to a Python environment (version 3.10.5). Detection of both ‘slow waves’ and ‘sleep spindles’ was performed according to previously described guidelines (28). For each channel, the densities of slow waves and sleep spindles were averaged across the channels for N2 and SWS phases.

#### Oxygen saturation and HR

During the participants’ registered sleep time, blood oxygen saturation (SpO_2_) and HR were continuously assessed using pulse oximetry (Nellcor PM10N, Medtronic, Minneapolis, USA). These data were used to calculate both average and minimal HR, the minimal SpO_2_, the average nocturnal SpO_2_ both over the entire night, as well as within 10% epochs of the night. Furthermore, variation in SpO_2_ was assessed through a coefficient of variation (CV) for every hour (CV_i_ = SD_i_ / mean_i_ with i = the hour of the night) in agreement with an earlier study (29). CV_i_ was calculated given earlier evidence that periodic breathing causes fluctuations in nocturnal oxygen saturation in preterm infants (30).

#### Resting measurements

All resting measurements were performed by the same trained researcher. Measurements were performed throughout a 10min time-window and with the participants being in supine position for at least 10min before the start of the first measurement. Data for all parameters are presented as the average values of the last minute.

#### Blood and tissue oxygenation status

Blood oxygen saturation (SpO_2_) was assessed using a pulse oximeter (Nellcor PM10N, Medtronic, Minneapolis, USA) with an infrared sensor placed ∼2 cm above the left eyebrow and with sampling frequency set at 2 Hz. Cerebral and skeletal muscle oxygenation status were assessed by means of tissue oxygenation index (cTOI and mTOI, resp.) using near infrared spectroscopy (NIRS). The probes of a NIRO-200 spectrometer (Hamamatsu, Japan) were attached ∼2 cm above the right eyebrow (cerebral oxygenation) and centrally on the belly of the right *m. vastus lateralis* (skeletal muscle oxygenation). A constant penetration depth of ∼2 cm into the muscle/brain tissue was ensured by inserting the emitter and detector probes in a dark-colored rubber spacer, maintaining a fixed interoptode distance of 4 cm. These spacers were attached to the participants using an elastic non-transparent bandage and double-sided adhesive tape to prevent displacement or interference from external light. Before each experimental session, the participants’ skin was shaved and cleaned to exclude any signal disturbance by hair or impurities. Moreover, exact inter-session replication was guaranteed by marking the contour lines of the rubber spacer on the skin. Participants were asked to preserve and refresh these marks during the washout period in order to maintain this position during the following sessions. After experimental data collection, NIRS data were preprocessed (Matlab R2023a, The Mathworks, Natick, MA) over 1-min long time chunks using a fourth-order Butterworth filter with a cut-off frequency of 0.05 Hz (24).

#### Heart rate and heart rate variability

Resting HR and RR-peak intervals were measured using a Polar H10 (Polar, Kempele, Finland). HRV was analyzed by using the percentage of adjacent NN intervals that differ by more than 50 ms (pNN50), the root mean square of successive differences (RMSSD), and absolute power of the high-frequency band (HF) using Kubios (Kubios HRV Standard 3.5.0, Kubios Oy, Kuopio, Finland).

#### Ventilatory gas exchange measurements

Indirect calorimetry (Cortex Metalyzer 3b, Leipzig, Germany) was used to measure breath-by-breath gas exchange data [*i.e.,* minute ventilation (VCE), oxygen uptake rate (VCO_2_) and carbon dioxide production rate (VCCO_2_)].

##### Exercise performance

After returning to normoxia on the second day, participants performed TT_30’_ which was preceded by a 15-min warm-up at 70% of LT on a cycling ergometer (Avantronic Cyclus II, Leipzig, Germany). Participants were instructed to achieve an average power output as high as possible and were allowed to voluntary change the applied workload every 5min in the initial 25min, and every 1min during the final 5min. The starting workload was determined during the familiarization sessions, where initial power in the first session was set at the LT and in the second as the average power output of the first TT_30’_. In turn, the average of the second TT_30’_ functioned as the starting workload of both experimental TT_30’_.

### Capillary blood, urine sampling and analyses

#### Capillary samples

Before breakfast (baseline, 0h), thirty min after every supplement ingestion (+5.5h, +9h and +11h and +13h), and immediately upon waking up on the second morning (+23h), a capillary blood sample was obtained for immediate determination of D-ß-hydroxybutyrate (GlucoMen Areo 2K-meter with ß-ketone sensor strips, A. Menarini Diagnostics, Firenze, Italy). In addition, 70 µl capillary blood was collected from a hyperemic earlobe into a capillary tube (safeCLINITUBE, Radiometer Medical ApS, Copenhagen, Denmark) before every resting measurement (baseline, +10h, +23h and +27h) and immediately before sleep (+13h). After immediate mixing for 10sec, samples were analyzed for acid-base balance, p50, and pCO_2_ (ABL90 FLEX analyzer, Radiometer Medical ApS, Copenhagen, Denmark).

#### Urine samples

Immediately before sleep time, participants emptied their bladder and nocturnal urine was collected until wake-up in urine flasks prepared with 10 mL hydrochloric acid. A small volume (∼10 mL) of urine was frozen (−80 °C) for subsequent analyses of adrenaline, noradrenaline, and dopamine using a commercially available ELISA kit (BA E-6600, LDN, Nordhorn, Germany) within 4 weeks after the final experimental session.

#### Statistical analyses

All statistical analyses were performed in GraphPad Prism version 10.1.2 (GraphPad Software, La Jolla, CA, USA). Prior to statistical testing, normal distribution of the data was evaluated and confirmed with the D’Agostino-Pearson normality test for SpO_2_ and HR during the night as well as power output and HR during TT_15’_. One way analysis of variance (ANOVA) was used to evaluate differences between conditions for measurements obtained at a single time point during each session. Data collected at multiple time points within a given experimental session were analyzed by means of a two-way repeated measures ANOVA. A Geisser-Greenhouse correction was applied whenever the assumption of sphericity was violated (Mauchly’s test, JASP version 0.18.1, JASP Team, Amsterdam, The Netherlands). In case of a significant main or interaction effect, *post-hoc* analyses were performed using Šidák’s correction. When applicable, reported p-values refer to these post-hoc analyses and otherwise, p-values for main effects were included. All data are presented as mean ± SD and statistical significance was defined as p < 0.05. An a-priori sample size calculation was performed based on the effect size of KE on sleep efficiency (i.e., primary outcome of the current study) that was derived from our previous study investigating the effect of KE during sleep in normoxia (25). Power analysis (G*Power version 3.1.9.7) indicated that a sample size of 6 per condition is required to establish a significant effect on sleep efficiency (effect size η_p_^2^: 0.45; α error: 0.05; power: 0.80; correlation among repeated measures: 0.5; nonsphericity correction: 1; ANOVA: repeated measures, within factors).

## RESULTS

### Blood βHB concentrations

A time x condition effect was observed for blood [βHB] (p < 0.001, Fig. 2). Baseline concentrations were low (∼0.4 mM) in all conditions, and concentrations remained low (∼0.4-0.5 mM) throughout the entire protocol in N_PL_ and H_PL_. Conversely, blood [ßHB] consistently increased to 2-3 mM (range: 1.4 to 4.7 mM) thirty min after ingestion of each KE supplement. Upon waking up the second day of the protocol, blood [βHB] had returned to baseline values in all conditions.

**Figure 2.**
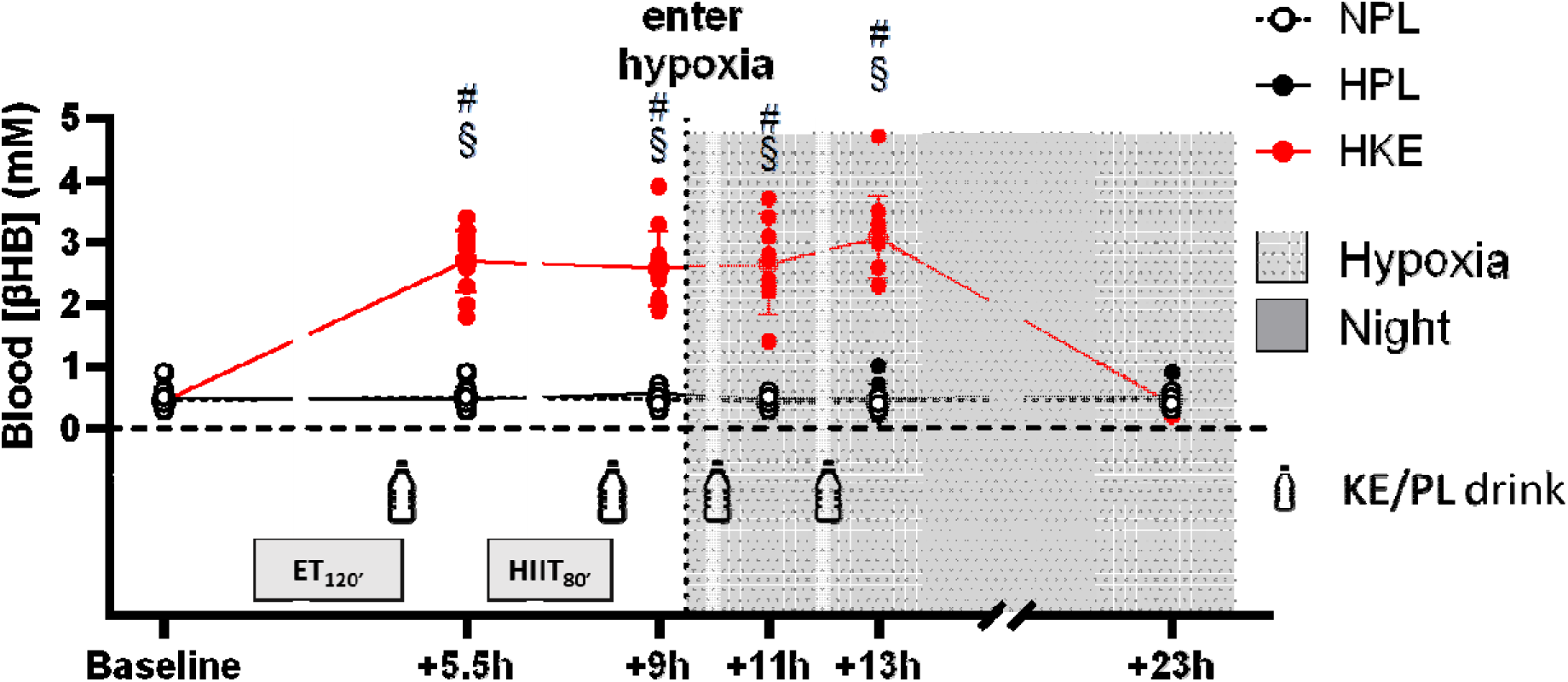
Blood D-β-hydroxybutyrate concentrations ([βHB]) are presented in response to ingestion of ketone ester (KE) or placebo (PL) supplements during experimental sessions including a night (dark grey zone) either in normoxia (N_PL_, black, open circles) or at a simulated altitude of 3,000m (grey dotted zone; H_PL_, black and H_KE_, red full circles). Blood [βHB] was assessed at baseline, 30min after the morning endurance training (ET_120’_; +5.5h) and the afternoon high intensity interval training (HIIT_80’_; +9h), as well as after 2h in hypoxia/normoxia (+11h), immediately before sleep (+13h) and upon waking up (+23h). Mean (line) ± SD (whiskers), as well as individual values are shown for n = 11. §, p < 0.05 *vs.* N_PL_; #, p < 0.05 *vs.* baseline for indicated condition.

### Sleep architecture

Time in bed was kept constant for a given subject across all conditions and was on average 512 ± 25min. Compared to N_PL_, H_PL_ decreased sleep efficiency by 3% (p = 0.037, Fig. 3a). This was mediated by a doubling (+ ∼17min) of WASO (p = 0.049, Fig. 3b), increased number of arousals (p = 0.027, Fig. 3c) while sleep onset latency (p = 0.316, Fig. 3d), and latencies of N2, SWS and REM phases remained unaffected (all p > 0.29, data not shown). The reduced sleep efficiency in H_PL_ *vs.* N_PL_ was accompanied by a ∼22% reduction in SWS (p = 0.002, Fig. 3e), and a ∼39% increase in N1 duration (p = 0.034, Fig. 3f), while N2 phase and REM sleep duration remained unaffected (p > 0.271, Fig. 3g-h, resp.). KE did not affect any of the hypoxia-induced dysregulations as all parameters were similar between H_PL_ and H_KE_ (all p > 0.340). Densities of both sleep spindles and slow waves during N2 (p = 0.082 and p = 0.307, resp.) and SWS (p = 0.752 and p = 0.782, resp.) were unaffected throughout the experimental sessions (data not shown).

**Figure 3.**
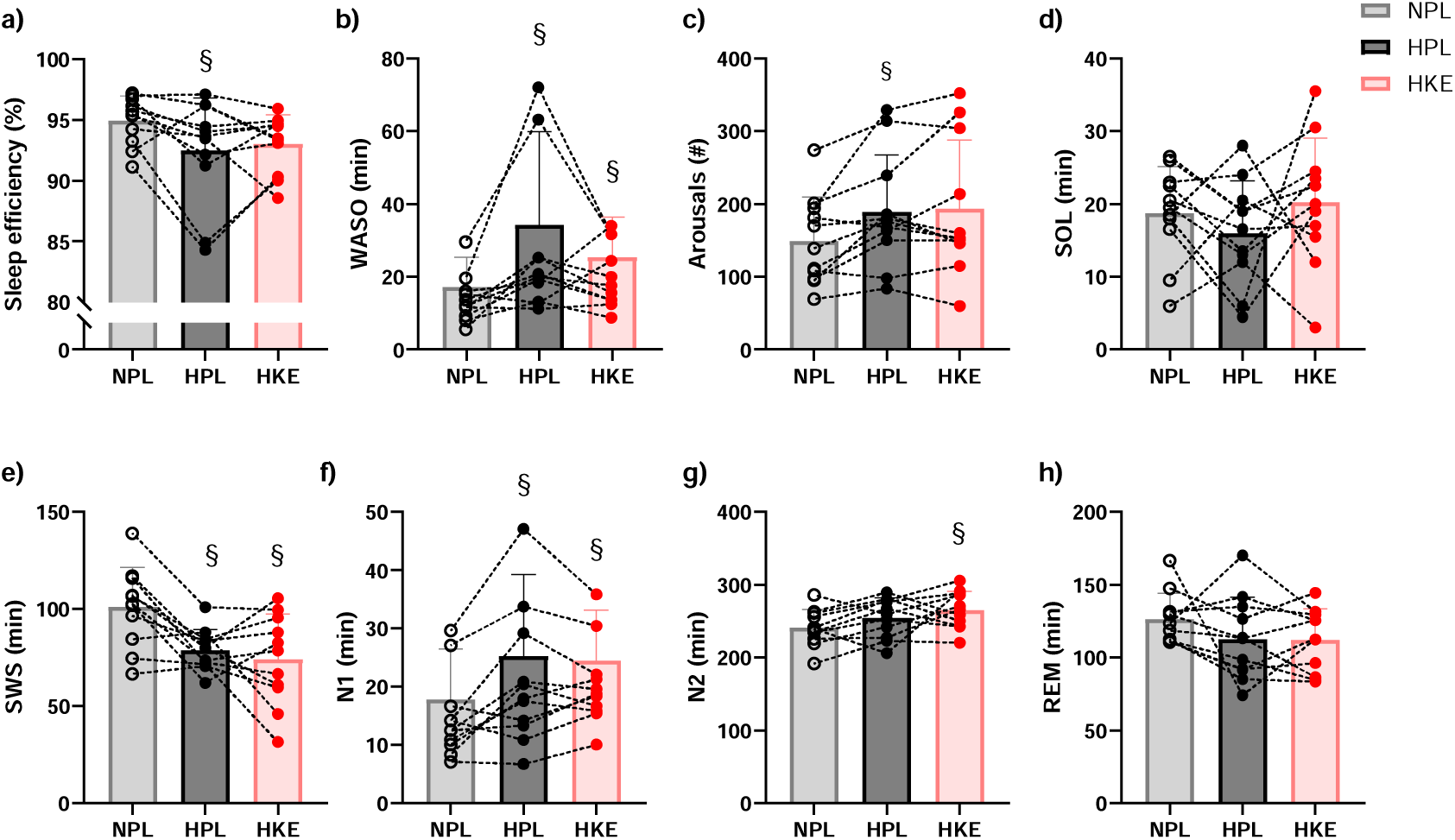
Data of polysomnographic (PSG) recordings are shown for participants that completed three experimental sessions in which they received either ketone ester (KE) or placebo (PL) supplements. They spent the night either in normoxia (N_PL_) or at a simulated altitude of 3,000m (H_PL_ and H_KE_). During the night, participants’ sleep was recorded using polysomnography (PSG). Mean (bar plots) ± SD (whiskers), as well as individual values are shown for a) sleep efficiency, b) wakefulness after sleep onset (WASO), c) the amount (#) of arousals per night, and d) sleep onset latency (SOL). Moreover, sleep stage durations are shown for e) slow wave sleep (SWS), f) N1 phase, g) N2 phase, and h) rapid eye movement (REM) sleep. Data are reported for 11 participants and datapoints are connected within each participant. §, p < 0.05 *vs.* N_PL_.

### Subjective sleep quality

A main effect was observed for the following questions in the St. Mary’s sleep questionnaire; *‘How deep was your sleep?’* (N_PL_: 5 ± 1, H_PL_: 4 ± 1, H_KE_: 4 ± 1; p = 0.007), *‘How many times did you wake up?’* (N_PL_: 3 ± 1, H_PL_: 4 ± 2, H_KE_: 4 ± 1; p = 0.007), *‘How clear-headed did you feel after getting up this morning?’* (N_PL_: 4 ± 1, H_PL_: 2 ± 1, H_KE_: 3 ± 1; p = < 0.001), *‘How satisfied were you with last night’s sleep?’* (N_PL_: 4 ± 1, H_PL_: 3 ± 1, H_KE_: 3 ± 1; p = 0.015). Post-hoc analyses indicated that, compared to N_PL_, participants in H_PL_ perceived that they slept less deep, woke up more times during the night, felt less clear-headed after getting up and were less satisfied with their night’s sleep. Except for a more clear-headed feeling upon waking up in H_KE_ (p = 0.012 *vs.* H_PL_), no differences were observed between H_PL_ and H_KE_. No effects were observed for the questions *‘How much sleep did you have last night?’* (N_PL_: 8 ± 1, H_PL_: 7 ± 1, H_KE_: 8 ± 1; p = 0.099), *‘How much sleep did you have during the day, yesterday?’* (N_PL_: 0 ± 0, H_PL_: 0 ± 0, H_KE_: 0 ± 0), *‘How well did you sleep last night?’*, (N_PL_: 4 ± 1, H_PL_: 3 ± 1, H_KE_: 3 ± 1; p = 0.051*), ‘Were you troubled by waking early and being unable to get off to sleep again?’*, (N_PL_: 1 ± 1, H_PL_: 0 ± 1, H_KE_: 1 ± 0; p = 0.123) and *‘How much difficulty did you have in getting off to sleep last night?’*, (N_PL_: 1 ± 1, H_PL_: 2 ± 1, H_KE_: 1 ± 0; p = 0.224)

### Nocturnal oxygen saturation and heart rate

At the start of the night, SpO_2_ was 9% lower in H_PL_ compared to N_PL_ (p < 0.001, Fig. 4a). Throughout the first half of the night, SpO_2_ gradually decreased in all conditions, reaching minimal SpO_2_ levels after ∼220min in bed or ∼44% of sleep for all conditions (p = 0.954 and p = 0.921, resp., data not shown). Due to both lower baseline values and a steeper reduction throughout the night in H_PL_ *vs.* N_PL_, minimal and average SpO_2_ were respectively 20 and 13% lower in H_PL_ compared to N_PL_ (p < 0.001, Fig. 4b-c). KE ingestion alleviated the gradual drop in SpO_2_ throughout the first part of the night (Fig. 4a). Hence, SpO_2_ after 40% of the night, as well as minimal SpO_2_ were both 4% higher in H_KE_ *vs.* H_PL_ (p = 0.029 and p = 0.048, Fig. 4a and Fig. 4b, resp.). Conversely, average nocturnal SpO_2_ was similar between H_KE_ and H_PL_ (p = 0.519 *vs.* H_PL_, Fig. 4c).

**Figure 4.**
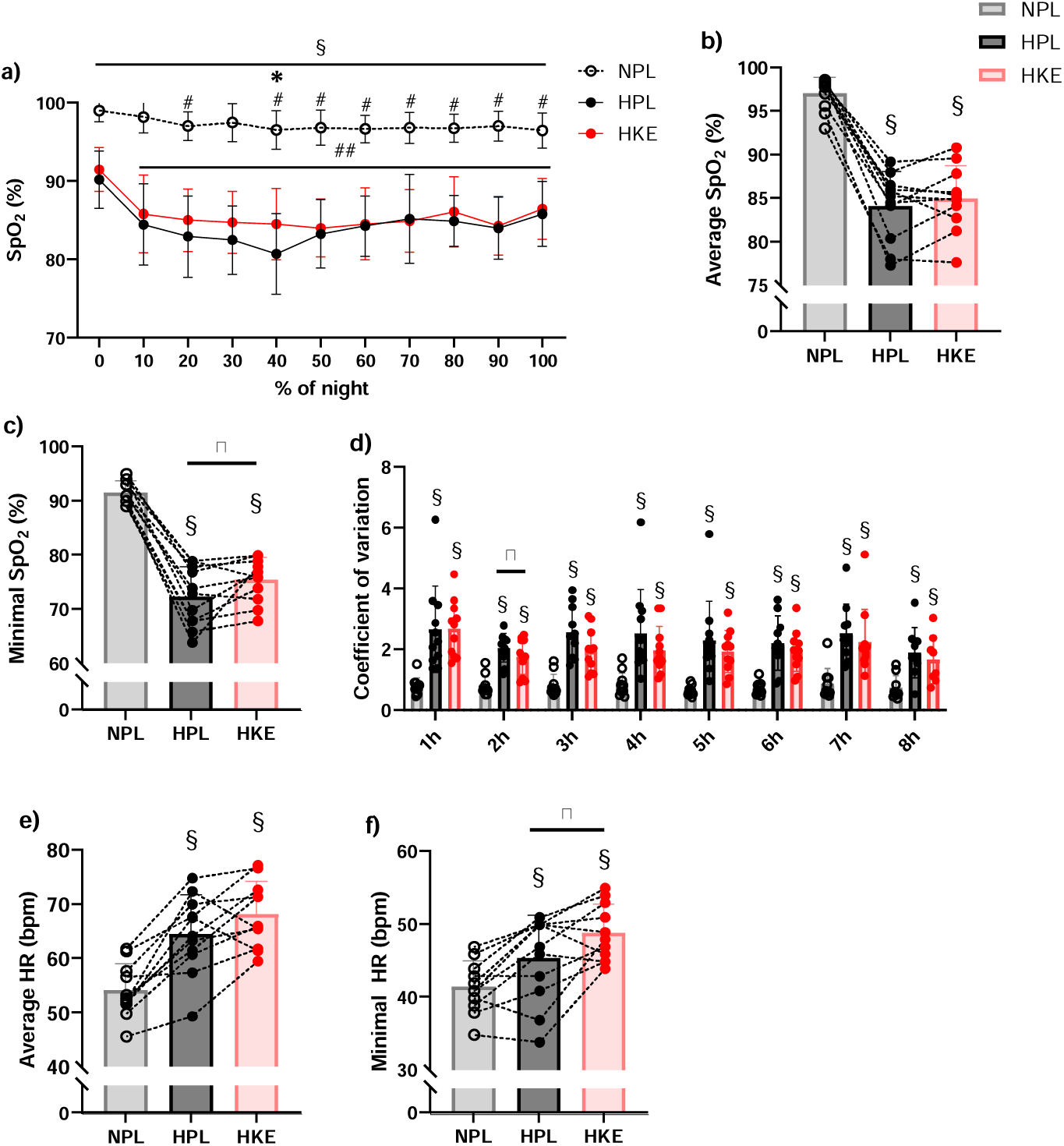
Nocturnal measurements of blood oxygen saturation (SpO_2_) and heart rate (HR) are shown for participants that completed three experimental sessions in which they received either ketone ester (KE) or placebo (PL) supplements. They spent the night either in normoxia (N_PL_) or at a simulated altitude of 3,000m (H_PL_ and H_KE_). Mean (circles) ± SD (whiskers) are shown for a) SpO_2_ throughout the percentual duration of the night (%). Mean (bar plots) ± SD (whiskers), as well as individual values are shown for b) average and c) minimal nocturnal _SpO2_ and d) coefficient of variance of SpO2 per hour of the night, as well as e) average and f) minimal nocturnal HR. Data are reported for 11 participants and individual values are connected within each participant. §, p < 0.05 *vs.* N_PL_; #, p < 0.05 *vs.* baseline; ##, p < 0.05 *vs.* baseline for H_PL_ and H_KE_; *, p < 0.05 for H_KE_ *vs.* H_PL_.

H_PL_ strongly increased the CV of SpO_2_ during all hours of the night compared to N_PL_ (p < 0.002, Fig. 4d). This increase in CV was attenuated by KE ingestion during the 2^nd^ hour of the night (p = 0.008 *vs.* H_PL_), but not at any of the other timepoints. Compared to N_PL_, H_PL_ increased minimal and average nocturnal HR by respectively ∼10% and ∼20% (p = 0.008 and p < 0.001 *vs.* N_PL_, resp, Fig. 4e-f). KE further increased minimal nocturnal HR by ∼8% (p = 0.019 for H_KE_ *vs.* H_PL_), and tended to further elevate average nocturnal HR (p = 0.079 for H_KE_ *vs.* H_PL_).

### Exercise performance

Mean power output during TT_30’_ was similar between all conditions at ∼285W [p = 0.454, confidence interval (CI): −5 to +9 W for N_PL_ *vs.* H_PL_ and −9 to +12 for H_KE_ *vs.* HPL, Fig. 5a,b). Also average HR during TT_30’_ (Fig. 5c) was similar between all conditions (p = 0.251). However, after 5 min of passive recovery, participants HR remained ∼6-9 bpm higher in H_PL_ and H_KE_ compared to N_PL_ (p < 0.001, Fig. 5d)

**Figure 5.**
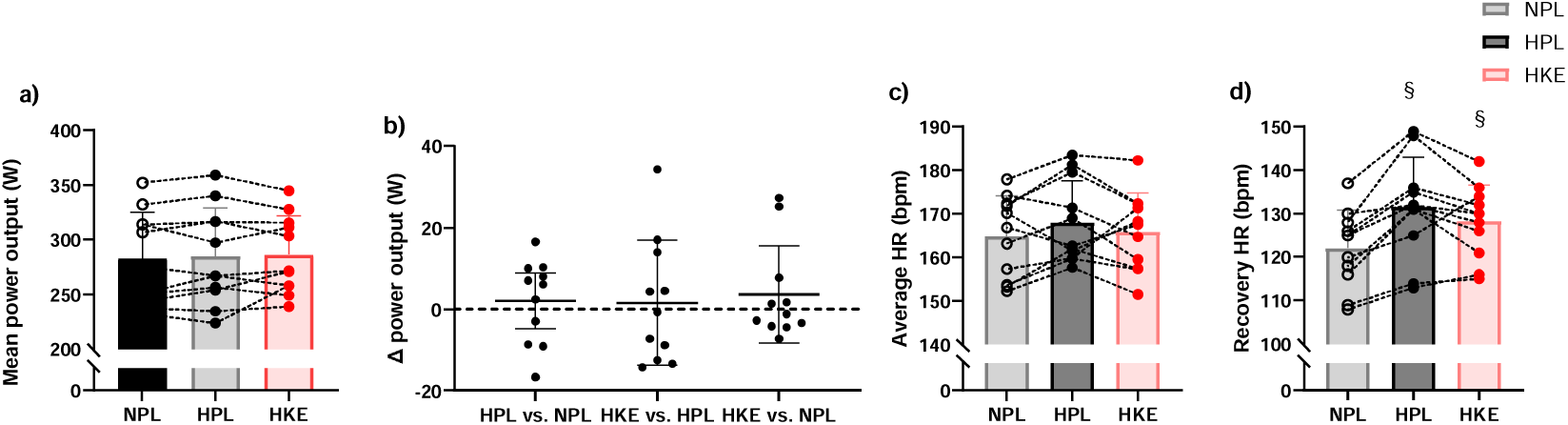
Exercise performance and heart rate data are shown for participants that completed three experimental sessions in which they received either ketone ester (KE) or placebo (PL) supplements during the first day. After spending the night in either in normoxia (N_PL_) or at a simulated altitude of 3,000m (H_PL_ and H_KE_), exercise performance was assessed by an all-out 30-min time trial (TT_30’_). Mean (bar plots) ± SD (a, c, d, whiskers) or mean (horizontal indication) ± 95% confidence intervals (b, whiskers), as well as individual values are shown for a) mean power output, b) the individual differences in mean power output between all experimental conditions and c) average heart rate (HR) during TT_30’_ and d) HR after 5min of passive recovery. Data are reported for 11 participants and individual values are connected within each participant §, p < 0.05 *vs.* N_PL._

### Resting measurements – oxygen status, heart rate and heart rate variability

A time x condition effect was detected for SpO_2_ (p < 0.001, Table 1). Compared to N_PL_, SpO_2_ dropped to a similar extent in H_PL_ and H_KE_ both at +10h (p < 0.001 *vs.* N_PL_) and +23h (p < 0.001 *vs.* N_PL_). Conversely, both cTOI and mTOI remained stable at respectively ∼73% and ∼67% in all experimental conditions (data not shown). For HR, a time x condition effect was found (p < 0.001, Table 1). Post-hoc analyses indicated that HR was ∼14% higher in H_PL_ *vs.* N_PL_ at +10h (p < 0.001), and even ∼20% higher at +23h (p < 0.001). KE ingestion further increased resting HR at +10h (p < 0.001 *vs.* H_PL_). An interaction effect was observed for all measured HRV indices (pNN50, p = 0.002; RMSSD, p < 0.001; HF, p = 0.005; Table 1). Compared to N_PL_, H_PL_ lowered pNN50, RMSSD and HF at +10h, while at +23h only pNN50 and RMSSD were decreased. KE did not alter the impact on RMSSD, but lowered pNN50 and HF at +23h compared to H_PL_.

**Table 1.** Effect of hypoxia (H_PL_ *vs.* N_PL_) and ketone ester (KE) ingestion (H_KE_ *vs.* H_PL_) on blood oxygen saturation (SpO_2_), heart rate (HR) and heart rate variability (HRV).

| | $N_{PL}$ | $H_{PL}$ | $H_{KE}$ |
| --- | --- | --- | --- |
| **$SpO_2$ (%)** |  |  |  |
| Baseline | $97.4 \pm 1.7$ | $98.5 \pm 1.5$ | $98.7 \pm 1.3$ |
| +10h | $98.2 \pm 1.1$ | $87 \pm 4$ §# | $88.5 \pm 4.5$ §# |
| +23h | $98.1 \pm 2.1$ | $89.2 \pm 2.6$ §# | $90.8 \pm 3.3$ §# |
| +27h | $98.2 \pm 1.7$ | $97.9 \pm 1.7$ | $98.7 \pm 1$ |
| **HR (bpm)** |  |  |  |
| Baseline | $60.1 \pm 6.1$ | $60.7 \pm 5.1$ | $57.6 \pm 6.1$ |
| +10h | $65.6 \pm 7.8$ # | $74.9 \pm 8.6$ §# | $84.8 \pm 7$ §#* |
| +23h | $51.4 \pm 6.3$ # | $61.3 \pm 9.7$ § | $63.6 \pm 8.4$ §# |
| +27h | $55.2 \pm 6.4$ # | $56.9 \pm 6.9$ | $58.2 \pm 7.8$ |
| **pNN50 (%)** |  |  |  |
| Baseline | $43.4 \pm 21.6$ | $45.2 \pm 24.8$ | $52.5 \pm 23.2$ |
| +10h | $34.4 \pm 23.5$ | $14.1 \pm 19.2$ §# | $4.2 \pm 7.6$ §# |
| +23h | $59.5 \pm 24.9$ | $38.3 \pm 21.3$ § | $22.3 \pm 17.1$ §*# |
| +27h | $47.4 \pm 23.5$ | $50.1 \pm 27.0$ | $51.9 \pm 23.2$ |
| **RMSSD (ms)** |  |  |  |
| Baseline | $75.4 \pm 41.7$ | $74.2 \pm 29.3$ | $86.7 \pm 37.4$ |
| +10h | $62.7 \pm 43.1$ | $32.1 \pm 22.2$ §# | $21.3 \pm 20.6$ §# |
| +23h | $94.2 \pm 38.4$ | $70.3 \pm 36.2$ § | $50.8 \pm 28.2$ §# |
| +27h | $88.9 \pm 38.1$ | $96.8 \pm 51.6$ | $87.8 \pm 38.5$ |
| **HF (log)** |  |  |  |
| Baseline | $7.42 \pm 1.00$ | $7.46 \pm 0.72$ | $7.41 \pm 1.00$ # |
| +10h | $7.04 \pm 1.00$ | $5.67 \pm 1.19$ §# | $4.75 \pm 1.56$ §# |
| +23h | $7.07 \pm 0.99$ | $7.14 \pm 0.91$ | $6.46 \pm 1.16$ * |
| +27h | $7.48 \pm 1.34$ | $7.5 \pm 1.25$ | $7.62 \pm 1.05$ |

During a randomized, cross-over protocol, participants received either ketone ester (KE) or placebo (PL) supplements and spent the night either at a simulated altitude of 3,000m ( $H_{PL}$ and $H_{KE}$ ) or in normoxia ( $N_{PL}$ ). Participants' blood oxygen saturation ( $SpO_2$ ), heart rate (HR) and heart rate variability (HRV) were assessed at baseline, after 2h in hypoxia/normoxia (+10h), immediately upon waking up in hypoxia/normoxia (+23h) and 2h after returning to normoxia (+27h). Mean values ± SD (n=11) are shown for $SpO_2$ , HR, the percentage of adjacent NN intervals that differ by more than 50 ms (pNN50), the root mean square of successive differences (RMSSD), and the absolute power of the high-frequency band (HF). §, $p < 0.05$ vs. $N_{PL}$ ; \*, $p < 0.05$ vs. $H_{PL}$ ; #, $p < 0.05$ vs. baseline.

### Resting measurements – acid-base balance, blood gasses and ventilatory parameters

Blood pH, p50, pCO_2_, as well as ventilatory parameters were similar between all conditions at baseline (Table 2). Compared to N_PL_, H_PL_ increased blood pH at +10h and +23h but not at +27h, while pCO_2_ was decreased at all timepoints. Conversely, p50 remained similar between N_PL_ and H_PL_ at all timepoints. Relative to H_PL_, H_KE_ caused a slight acidosis at +10h which was accompanied by a KE-induced reduction in pCO_2_, and an increase in p50. Each of these KE-induced alterations were again normalized at +23h and +27h. Irrespective of the experimental condition, VCE was similar at baseline (11.5 ± 0.2 L.min^−1^), increased at +10h (13.1 ± 0.2 L.min^−1^, p < 0.001 *vs.* baseline) and decreased at +23h (9.9 ± 0.4 L.min^−1^, p = 0.001 *vs.* baseline). VCO_2_ was similar at baseline (0.161 ± 0.014 L.min^−1^) and increased at +10h (0.236 ± 0.009 L.min^−1^, p = 0.004 *vs.* baseline). Also VCCO_2_ was similar at baseline (0.182 ± 0.012 L.min^−1^) yet decreased at +27h (0.0125 ± 0.017 L.min^−1^, p = 0.018 *vs.* baseline).

**Table 2.** Effect of hypoxia (H_PL_ *vs.* N_PL_) and ketone ester (KE) ingestion (H_KE_ *vs.* H_PL_) on blood acid base balance and capillary blood gases.

| | $N_{PL}$ | $H_{PL}$ | $H_{KE}$ |
| --- | --- | --- | --- |
| **pH** |  |  |  |
| Baseline | $7.409 \pm 0.016$ | $7.412 \pm 0.013$ | $7.406 \pm 0.011$ |
| +10h | $7.419 \pm 0.021$ | $7.438 \pm 0.017$ §# | $7.395 \pm 0.016$ §#* |
| +13h | $7.414 \pm 0.014$ | $7.433 \pm 0.018$ §# | $7.412 \pm 0.027$ |
| +23h | $7.421 \pm 0.013$ # | $7.451 \pm 0.026$ §# | $7.457 \pm 0.009$ §# |
| +27h | $7.417 \pm 0.015$ | $7.431 \pm 0.016$ # | $7.432 \pm 0.018$ # |
| **p50 (mmHg)** |  |  |  |
| Baseline | $26.2 \pm 0.5$ | $25.9 \pm 0.7$ | $25.9 \pm 0.9$ |
| +10h | $25.1 \pm 1$ # | $25.9 \pm 0.7$ | $26.8 \pm 1.3$ §* |
| +13h | $25.8 \pm 0.9$ | $25.8 \pm 1$ | $25.5 \pm 1.4$ |
| +23h | $25.3 \pm 1.3$ | $25 \pm 1$ | $24.4 \pm 1$ §# |
| +27h | $25.6 \pm 1.1$ | $25.7 \pm 0.6$ | $25.1 \pm 1.2$ |
| **pCO<sub>2</sub> (mmHg)** |  |  |  |
| Baseline | $41.2 \pm 2.2$ | $42 \pm 1.8$ | $41.8 \pm 1.7$ |
| +10h | $42.4 \pm 1.1$ | $40.9 \pm 1.4$ § | $38 \pm 1.2$ §* |
| +13h | $42.4 \pm 1.6$ | $40 \pm 2$ § | $37.3 \pm 2$ §* |
| +23h | $41.5 \pm 2.2$ | $37 \pm 2.1$ § | $36.7 \pm 1.5$ § |
| +27h | $42.3 \pm 1.7$ | $39.4 \pm 1.6$ § | $39 \pm 1.9$ § |

During a randomized, cross-over protocol, participants received either ketone ester (KE) or placebo (PL) supplements and spent the night either at a simulated altitude of 3,000m ( $H_{PL}$ and $H_{KE}$ ) or in normoxia ( $N_{PL}$ ). Participants' capillary blood was analyzed baseline, after 2h in hypoxia/normoxia (+10h), immediately before sleep (+13h) and upon waking up in hypoxia/normoxia (+23h) and 2h after returning to normoxia (+27h). Capillary blood pH, p50 and pCO<sub>2</sub> were evaluated. §, $p < 0.05$ vs. $N_{PL}$ ; \*, $p < 0.05$ vs. $H_{PL}$ ; #, $p < 0.05$ vs. baseline.

### Nocturnal catecholamine excretion

Nocturnal urine production was similar in all conditions (N_PL_: 679 ± 375 mL, H_PL_: 626 ± 223 mL, H_KE_: 685 ± 321 mL, p = 0.647). Also nocturnal adrenaline (N_PL_: 5.86 ± 3.66 nmol, H_PL_: 6.93 ± 4.16 nmol, H_KE_: 6.60 ± 4.14 nmol, p = 0.305), noradrenaline (N_PL_: 52.5 ± 22.2 nmol, H_PL_: 50.3 ± 10.8 nmol, H_KE_: 56.7 ± 12.4 nmol, p = 0.608) and dopamine (N_PL_: 911 ± 305 nmol, H_PL_: 795 ± 229 nmol, H_KE_: 956 ± 350 nmol, p = 0.324) excretions were unaffected by the different conditions.

## DISCUSSION

Sleeping at (simulated) altitude is increasingly common in athletes either as part of altitude training camps or due to competitions at such altitudes. Earlier studies indicated that, starting from ∼2,000m, this typically causes an acute reduction in sleep quality (2–10) thereby potentially impairing exercise recovery and readiness to perform. We recently observed that KE ingestion counteracts sleep disruptions induced by late-evening exercise under normoxic conditions (25). Furthermore, KE has also been shown to attenuate hypoxemia, which is considered the primary mechanism underlying hypoxia-induced sleep disturbances (23, 24). Therefore, we investigated whether KE ingestion could counteract sleep disruptions induced by sleeping at altitude (i.e., 3,000m), and impact next day endurance exercise performance. Sleeping at altitude impaired sleep quality as evidenced by a 3% reduction in sleep efficiency, a doubling of WASO and a ∼20% reduction in slow wave sleep. These sleep disturbances did not affect performance during a 30-min time-trial the next day, but reduced subsequent heart rate recovery. KE slightly alleviated nocturnal hypoxemia but was unable to negate these sleep disruptions, nor impacted next day exercise performance or subsequent heart rate recovery.

Based on the available literature, sleeping at 3,000m altitude is expected to increase WASO (3–7, 9) and potentially decrease slow wave sleep (2, 5, 6, 9, 13), while REM sleep and sleep efficiency generally remain unaffected up to altitudes of ∼4,000m (2, 4, 6, 7, 11–13). Although our results on WASO, slow wave sleep and REM are in line with available literature, we observed that even lower simulated altitudes (i.e. 3,000 m) can already elicit a drop in sleep efficiency. As sleep efficiency depicts the time spent asleep over the time spent in bed, with the latter standardized over all sessions, this drop was established by a decreased time asleep through a doubling in WASO. The divergent *vs.* convergent effects on respectively SE and WASO between our study and earlier studies likely resulted from the fact that we, in contrast to most of the earlier studies, standardized the total time in bed. This is supported by two earlier studies showing that under free-living conditions the observed increase in nocturnal wakefulness upon altitude is at least in part compensated by a ∼5% increase in time in bed (6, 7). Nevertheless, this increase in time in bed failed to reach statistical significance in both studies.

Increased awakenings upon hypoxia are generally attributed to a hypoxia-induced periodic breathing pattern (5, 31). This phenomenon almost exclusively occurs during NREM sleep and especially during SWS, and causes short awakenings or arousals and disruption of SWS. Our observations indicating that hypoxia decreases SWS but not REM sleep, together with an increased amount of arousals, thus indirectly endorse the important role of periodic breathing in hypoxic sleep impairment. This is further supported by our observation that hypoxia drastically increased the periodic variation of SpO_2_ during the night, which is a central hallmark of periodic breathing (29).

Sleep disturbances can evoke a myriad of negative effects on processes implicated in exercise performance, training adaptation, and general health. This is evidenced by studies showing that large reductions in sleep quantity (reductions of 2-3 h.night^−1^ for 1 to 5days) for instance increase heart rate during submaximal exercise (14), suppress myofibrillar and sarcoplasmic protein synthesis rates (15, 16), and impair mitochondrial respiratory function (16) and insulin sensitivity (17). Although some studies reported a concomitant drop in both sprint and endurance cycling performance (19, 20), even such large sleep disruptions do not always evoke impairments in exercise performance (14, 18). However, to our knowledge, no data is available on the impact of more subtle sleep disturbances such as those observed in our study.

The minor sleep disturbances induced by hypoxia in our study did not affect TT_30’_ performance, nor altered resting heart rate, heart rate variability or respiratory parameters the day after. However, the (disrupted) night in hypoxia compromised heart rate recovery after TT_30’_ as evidenced by the ∼10 bpm higher heart rate in H_PL_ *vs.* N_PL_ five min after completion of TT_30’_. A reduced or delayed heart rate recovery reflects an impaired vagal activity (32), a strong predictor for morbidity (33), which is generally associated with cardiovascular disease (34) and overtraining (35). This impaired heart rate recovery upon hypoxia may be related to the hypoxia-induced reduction in SWS. Indeed, selective SWS suppression was previously found to disrupt sympathovagal balance by decreasing vagal tone (36) and stimulation of slow waves during sleep improved next day cardiac function, as evidenced through an improved left-ventricular systolic function (37, 38). This indicates that although exercise performance was not affected after one disrupted night, more subtle changes are present which may in the longer term impact exercise recovery, performance or general (cardiovascular) health.

In contrast to our hypothesis, KE ingestion did not counteract the hypoxia-induced sleep dysregulations. This conflicts with our earlier data showing that KE intake negated the sleep disruptions induced by strenuous late evening exercise under normoxic conditions (25). A potential explanation for these contrasting observations are the disparate effects of strenuous late evening exercise *vs.* hypoxia on sleep architecture. Both conditions caused a decrease in sleep efficiency and an increase in WASO. However, while strenuous late evening exercise decreased REM sleep without affecting NREM sleep, hypoxia did not affect REM sleep but decreased SWS. This suggests that ketosis may only beneficially impact sleep when REM is disrupted. This is supported by an earlier study showing that a ketogenic diet improved REM sleep in children with epilepsy (39), a condition typically associated with reduced REM sleep (40). More recent data collected by our group during a 7-week exercise training program confirms this hypothesis as KE only increased REM sleep relative to a placebo group whenever REM sleep dropped below baseline values (Robberechts et al., unpublished observations). We previously suggested that the ability of KE to counteract reductions in REM sleep was mediated through an increased dopamine signaling following KE ingestion (41, 42), as dopamine plays an important role in the transition from NREM to REM sleep (43). However, in contrast to our earlier studies, KE intake did not affect nocturnal dopamine excretion in the current study.

An important factor in compromised hypoxic sleep is hypoxemia, and subsequent periodic breathing. As described above, periodic breathing induces periods of hypopnea and apnea causing short awakenings especially during SWS (12). As minimal SpO_2_ values are closely related to the apneic part of a periodic breathing cycle (44), the observed KE-induced increase (∼+3%) in minimal SpO_2_ and SpO_2_ at 40% of sleep duration together with the diminished variance in nocturnal SpO_2_ after 2h of sleep suggest a slightly tempered periodic breathing behavior after KE ingestion. However, this effect was likely to small to translate in sleep improvements. As nocturnal respiration was not recorded in the current study, further research is required to identify if KE may impact periodic breathing.

On the other hand, hypoxemia activates a wide array of adaptive responses, amongst others establishing the respiratory alkalosis observed in H_PL_, causing a left-shift of the oxyhemoglobin dissociation curve (ODC) and thus favoring a higher SpO_2_ for a given pO_2_. Ingestion of KE, contrarily, established a relative metabolic acidosis in agreement with earlier data (23, 24), thereby pushing a right-shift of the ODC as evidenced by the observed increase in p50. Nevertheless, average nocturnal SpO_2_ values were similar after ingestion of KE compared to PL and only after 40% of sleep duration, SpO_2_ was higher in KE. This can at least partly be explained by the higher capillary pO_2_ values that were observed immediately before sleep, i.e. after 5h in hypoxia, in H_KE_ (56.8 ± 3.4 mmHg) *vs.* H_PL_ (53.3 ± 4.0 mmHg) thereby compensating the right shift of the OCD. It should however be highlighted that pO_2_ values were obtained using a capillary sampling method, which might underestimate actual arterial pO_2_.

As indicated above, KE increased SpO_2_ at 40% of the night, as well as minimal nocturnal SpO_2_, but did not impact SpO_2_ at any other timepoint. This is most likely related either to (i) the extent of hypoxemia or (ii) to the specific duration of ketosis or hypoxic exposure. This is supported by our two earlier studies showing that KE attenuated hypoxemia only after ∼3-4h of hypoxic exposure and ketosis, and whenever SpO_2_ values were below ∼82% (23, 24). These requirements were fulfilled at 40% of the night, but not at the other timepoints where we measured SpO_2_. For instance, at +10h participants were in ketosis but had only resided in hypoxia for 2h, while at +23h, participants had resided in hypoxia for 14h however were no longer in ketosis. Furthermore, at the start of the night SpO_2_ values were around 90% while during the final part of the night participants were most likely no longer in ketosis (25).

Besides hypoxemia, ketone bodies may also affect sleep by altering sympathetic nervous activity. In line with literature (45, 46), hypoxia caused an increase in both resting and nocturnal heart rate, as well as a decrease in all HRV parameters on the first evening of the protocol thereby suggesting a relative dominance of the sympathetic nervous system. Upon waking up however, HF values had recovered but RMSSD and pNN50 values remained depressed. While RMSSD and pNN50 are measures for overall HRV, HF isolates the high-frequency component (0.12-0.4 Hz) of the spectrum and entails information on ventilation and respiratory sinus arrhythmias (RSA), a phenomenon explaining fluctuation of HR and HRV together with natural breathing (47–49). As natural breathing is disrupted in hypoxia, both through periodic breathing and consistent hyperventilation, HF was indeed expected to increase (49). While SWS is associated with low heart rate variability in normoxia (50), sympathetic nerve activity is even higher during REM sleep compared to being awake (51). Although the exact relationship between sleep and sympathetic activity remains unclear, it cannot be ruled out that this increased sympathetic dominance is linked to the hypoxia-induced disruption of SWS. However, whether this is either a cause or a consequence remains to be identified.

Despite unaffected nocturnal adrenaline and noradrenaline concentrations, KE lowered HRV indices pNN50 and RMSSD upon waking up and increased nocturnal HR. This indicates that, in line with earlier studies performed both in normoxia (25, 52, 53) and hypoxia (23), KE ingestion further promoted sympathetic dominance. In this context, through manipulation of sympathetic activity, ketone bodies may only potentially benefit sleep quality when REM sleep, and not SWS, is disrupted.

In conclusion, we observed that a single night of hypoxic exposure (∼3,000m altitude) has a detrimental impact on sleep quality, through reduced sleep efficiency, SWS, and increased WASO. While this did not impact next day exercise performance, it hampered post-exercise heart rate recovery suggesting that sleeping at (simulated) altitude has minor implications for athlete training management. Contrary to our expectations, ketone ingestion did not mitigate any of these effects. However, we confirmed earlier data showing that KE can alleviate hypoxemia whenever SpO_2_ values drop below ∼82% and whenever subjects are at least for 3-4h in ketosis. Taken together with our earlier observations, ingestion of ketones as a strategy to improve sleep and recovery appears effective when REM sleep is disrupted (e.g., through late-evening exercise) whereas disruption of NREM sleep (through hypoxic exposure) remains unaffected.

## Data Availability

All data produced in the present study are available upon reasonable request to the authors.

## Competing interests

The authors declare that they have no competing interests.

## Author contributions

All experiments were performed within the Exercise Physiology Research Group and the Bakala Academy-Athletic Performance Center at the KU Leuven, Belgium. Conception and design of the study: MS, TD and CP. Data collection and/or data analyses: MS, DT, WL, RR, MR, TD and CP. Interpretation of the data and manuscript drafting: MS and CP. All authors critically revised the manuscript and approved the final version of the manuscript.

## Funding

This research was supported by the Research Foundation – Flanders (FWO Weave, research grant G073522N) and Slovene Research Agency grant (N5-0247). CP is supported by an FWO senior postdoctoral research grant (12B0E24N).

## Acknowledgements

The authors wish to thank all participants for their dedicated cooperation in this study. We also thank Ms. Monique Ramaekers for skillful assistance during the experimental trials.

## Notes

### Competing Interest Statement

The authors have declared no competing interest.

### Clinical Trial

NCT06060093

### Author Declarations

Ethics Committee Research (EC Research) of University Hospital Leuven (UZ Leuven) gave ethical approval for this work.

